# Development and Pilot Testing of a Mobile App Psychosocial Intervention for Psychological Distress in Individuals with Glaucoma

**DOI:** 10.64898/2026.05.20.26353674

**Authors:** Hannah M Fisher, Natalie A Chou, Margaret Falkovic, Heather Parnell, Christina Makarushka, Laura J Fish, Jennifer Plumb Vilardaga, Felipe A Medeiros, Tamara J Somers, Kelly W Muir, Samuel I Berchuck

## Abstract

**Objective:** To assess the feasibility and acceptability of VISON-ACT, a standalone, mobile app psychosocial intervention for psychological distress in individuals with primary open-angle glaucoma (POAG).

**Design:** Single-arm pilot.

**Participants:** Patients (N=28) with a diagnosis of POAG, self-reporting at least mild (≥3) distress on the 4-item Patient Health Questionnaire, were recruited from the Duke Eye Center between April 2025-December 2025.

**Methods:** Patients (n=28) were consented and completed a baseline (A1) self-report assessment. VISION-ACT was comprised of 6 weekly modules. Follow-up self-report assessments occurred at post- (A2) and 1-month post-intervention (A3) and included measures of psychological distress, vision and health-related quality of life, psychological flexibility, disease acceptance, self-efficacy for symptom management, mindfulness, and social support. Participants were invited to complete an exit interview at 1-month post-intervention to gather qualitative feedback on the VISION-ACT protocol. Descriptive statistics were used to assess feasibility and acceptability metrics and patterns of pre-post change on patient reported outcomes were explored with linear mixed mdels using R Statistical Software.

**Main Outcome Measures:** Feasibility (target accrual (n=25) in 12 months, <20% attrition at post-intervention); Acceptability (≥75% reporting use of VISION-ACT skills or ideas at post-intervention, ≥80% reporting M≥3.00/4.00 at post-intervention on the Client Satisfaction Questionnaire); Psychological Distress (Hospital Anxiety and Depression Scale [HADS], Subjective Units of Distress Scale [SUDS]).

**Results:** VISION-ACT was highly feasible; accrual target was surpassed (N=28) in 6 months, and attrition was low (3.85%) at post-intervention (A2). Acceptability was strong with 100% of participants reporting use of VISION-ACT skills or ideas at A2 and M=3.27/4.00 intervention satisfaction. Adherence was remarkable with 88.5% of participants completing all six VISION-ACT modules. Pre-post change patterns were in the expected direction for psychological distress (HADS A1 M=13.88, A2 M=11.21; SUDS A1 M=35.54, A2 M=26.46) and all other patient-reported outcomes across baseline, post- and 1-month post-intervention assessments. Data on participant perspectives highlighted valuable aspects of VISION-ACT, and areas for refinement.

**Conclusions:** Robust feasibility and acceptability data seen here provide support a fully-powered, randomized trial to evaluate the efficacy of VISION-ACT for reducing psychological distress and improving related patient-reported and clinical outcomes.

## INTRODUCTION

Glaucoma is the leading cause of irreversible vision loss worldwide and is projected to affect more than 100 million individuals by 2040.^1^ In the United States alone, more than 80,000 individuals experience glaucoma-related vision loss each year.^2,3^ Beyond its visual consequences, glaucoma imposes a substantial psychological burden, as patients must cope with the ongoing threat of progressive and permanent vision loss.^4,5^ Symptoms of anxiety and depression are common, affecting up to 25-30% of patients with glaucoma.^6,7^ Importantly, psychological distress (i.e., anxiety, depressive symptoms) has been linked to poorer adherence to treatment^8-11^ and follow-up,^12,13^ lower vision-related quality of life (QoL),^14,15^ and faster disease progression.^6^

Despite its high prevalence and clinical relevance, psychological distress is infrequently assessed and addressed in routine glaucoma care. Prior work, including our own, has shown that patients with glaucoma are receptive to screening for distress and that screening tools suitable for clinical use are readily available.^16,17^ However, systematic screening has not been widely adopted, and even more critically, psychological distress in the glaucoma context remains undertreated.^18^ In review studies, education-based and supportive interventions for patients with glaucoma, including mindfulness-based approaches and relaxation training, have been associated with improvements in intraocular pressure (IOP) and QoL, though this literature is nascent.^19,20^ Still, findings highlight the potential benefit of non-pharmacologic, psychosocial interventions in this patient population, as has been seen in other chronic disease groups (e.g., cancer).^21-25^ To date though, existing research has not rigorously examined whether an evidence-based, psychosocial intervention teaching multiple coping skills might be feasible and acceptable, and helpful for reducing psychological distress in patients with glaucoma.

Acceptance and Commitment Therapy (ACT) is cognitive-behavioral approach that teaches mindfulness and acceptance skills, and provides guidance for identifying values (e.g., being kind, being creative) and engaging in activities that are in line with those values (e.g., time with family, participation in hobbies).^26^ ACT may be particularly useful for patients with glaucoma because of its emphasis on psychological flexibility; that is, acceptance of uncontrollable situations (e.g., a glaucoma diagnosis and progression) while remaining committed to value-driven activities even when distress persists due to an ongoing stressor.^26,27^ A large and growing body of evidence supports the efficacy of ACT for improving psychological distress and related physical symptom and QoL outcomes in patients with medical conditions such as chronic pain and cancer.^23-25^ Notably, the application of ACT to the glaucoma experience has not yet been explored. This is a missed opportunity to leverage an evidence-based, psychosocial intervention likely well-suited to the challenging psychological burden of glaucoma.

To address this gap, our team of glaucoma physicians and clinical health psychologists with expertise in ACT approaches for managing chronic disease developed VISION-ACT. VISION-ACT is a standalone mobile app intervention comprised of six modules focused on central ACT domains of mindfulness, acceptance, and values clarification and engagement.^26^ The initial VISION-ACT protocol was iteratively refined through semi-structured interviews wherein glaucoma patients and clinicians (e.g., glaucoma physician, technician) were asked to reflect on the lived experience of glaucoma and preferences for intervention content, delivery, and features (see Supplementary Table 1 and 2 for summarizations). Rich qualitative data guided application of traditional ACT skills to the glaucoma context. For instance, key mindfulness and acceptance strategies (e.g., Grounding in Simple Awareness, Slow Belly Breaths) are recommended for tolerating commonly cited emotions during glaucoma, such as worry and fear. Likewise, Leaves on Stream, a cognitive diffusion technique, is encouraged for coping with distressing thoughts about glaucoma progression (e.g., “My vision will get worse,” “I will lose my independence”). Finally, suggestions for enhancing the user experience of the mobile app (e.g., larger font, high contrast visuals, inclusion of education materials) were instrumental to ensuring VISION-ACT was optimized for the unique needs of this population.

**Table 1.**
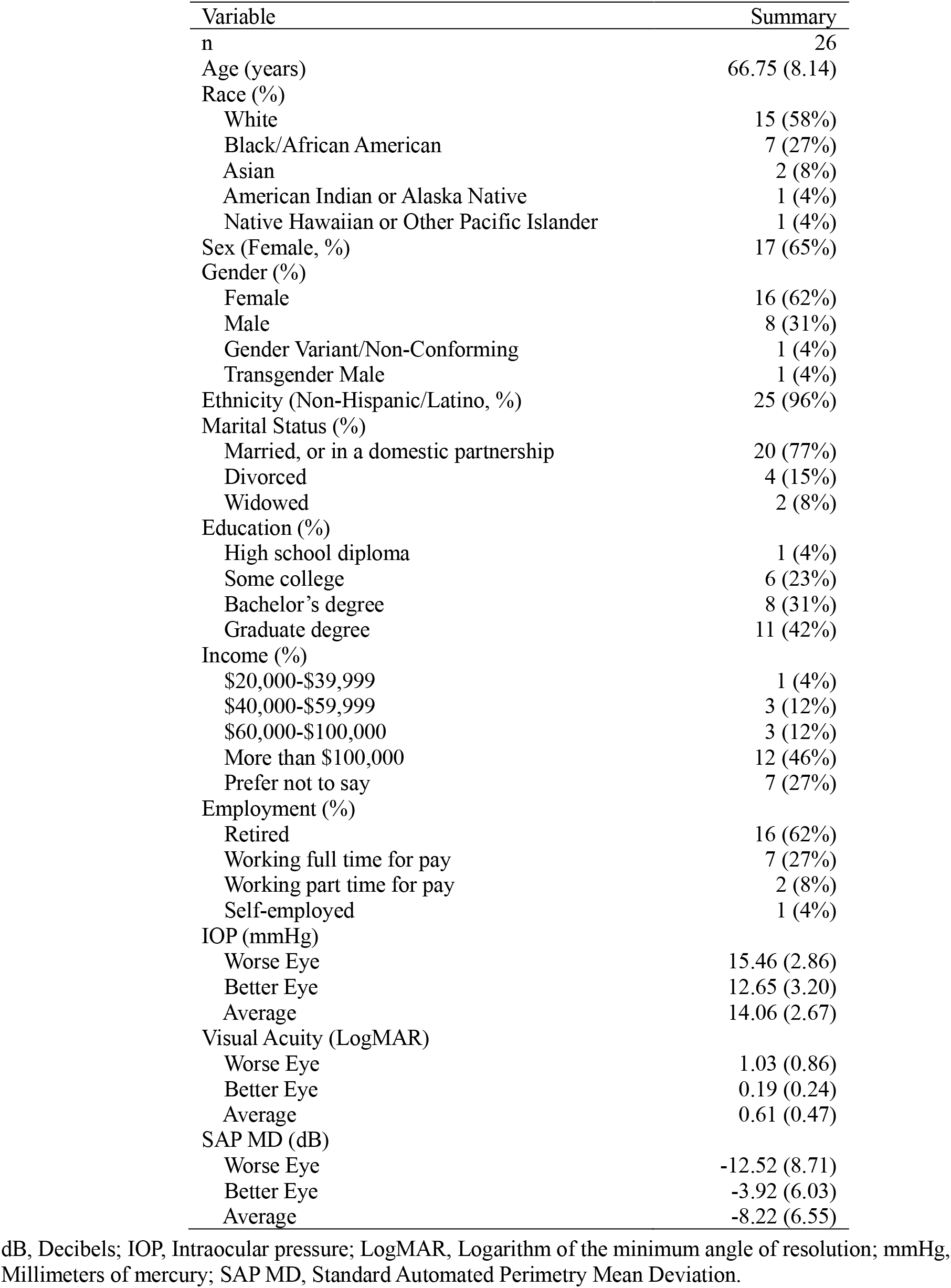
Summary statistics for VISION-ACT cohort presented as mean and standard deviation of continuous variables and counts and percentages for categorical variables. Eye specific variables are reported for the worse, better eyes, and averaged between the two eyes. Summaries are only shown for participants who started the intervention (n=26).

| Variable | Summary |
| --- | --- |
| n | 26 |
| Age (years) | 66.75 (8.14) |
| Race (%) |  |
| White | 15 (58%) |
| Black/African American | 7 (27%) |
| Asian | 2 (8%) |
| American Indian or Alaska Native | 1 (4%) |
| Native Hawaiian or Other Pacific Islander | 1 (4%) |
| Sex (Female, %) | 17 (65%) |
| Gender (%) |  |
| Female | 16 (62%) |
| Male | 8 (31%) |
| Gender Variant/Non-Conforming | 1 (4%) |
| Transgender Male | 1 (4%) |
| Ethnicity (Non-Hispanic/Latino, %) | 25 (96%) |
| Marital Status (%) |  |
| Married, or in a domestic partnership | 20 (77%) |
| Divorced | 4 (15%) |
| Widowed | 2 (8%) |
| Education (%) |  |
| High school diploma | 1 (4%) |
| Some college | 6 (23%) |
| Bachelor's degree | 8 (31%) |
| Graduate degree | 11 (42%) |
| Income (%) |  |
| \$20,000-\$39,999 | 1 (4%) |
| \$40,000-\$59,999 | 3 (12%) |
| \$60,000-\$100,000 | 3 (12%) |
| More than \$100,000 | 12 (46%) |
| Prefer not to say | 7 (27%) |
| Employment (%) |  |
| Retired | 16 (62%) |
| Working full time for pay | 7 (27%) |
| Working part time for pay | 2 (8%) |
| Self-employed | 1 (4%) |
| IOP (mmHg) |  |
| Worse Eye | 15.46 (2.86) |
| Better Eye | 12.65 (3.20) |
| Average | 14.06 (2.67) |
| Visual Acuity (LogMAR) |  |
| Worse Eye | 1.03 (0.86) |
| Better Eye | 0.19 (0.24) |
| Average | 0.61 (0.47) |
| SAP MD (dB) |  |
| Worse Eye | -12.52 (8.71) |
| Better Eye | -3.92 (6.03) |
| Average | -8.22 (6.55) |

dB, Decibels; IOP, Intraocular pressure; LogMAR, Logarithm of the minimum angle of resolution; mmHg, Millimeters of mercury; SAP MD, Standard Automated Perimetry Mean Deviation.

Early-stage work that follows established models of behavioral intervention development (e.g., NIH Stage Model)^28^ to carefully adapt ACT for glaucoma is noticeably absent from extant literature. To our knowledge, VISION-ACT is the first, and only, ACT-based, psychosocial intervention developed to specifically address psychological distress in patients with glaucoma. Pilot testing is indicated to assess the feasibility and acceptability of VISION-ACT, particularly with respect to its mobile app delivery format. The few education-based^29,30^ and supportive^31^ interventions for patients with glaucoma that do exist are primarily therapist-led and have not been developed with scalability in mind since access to behavioral care is limited by a shortage of trained behavioral health providers in the medical context.^32^ Mobile health (mHealth) offers a promising strategy to expand access to evidence-based psychosocial interventions targeting disease symptoms (e.g., psychological distress) that can be scalable. mHealth technologies, specifically mobile apps, have been shown to be a feasible and acceptable delivery format for chronic disease symptom management interventions.^33-35^ Moreover, several randomized trials examining entirely asynchroncous, mobile app-delivered psychosocial interventions show they are often non-inferior to in-person protocols for reducing psychological distress and improving QoL across a range of chronic disease populations.^33,36-38^ In glaucoma, however, existing mobile apps have focused primarily on disease monitoring and treatment adherence, with little attention to psychological outcomes.^39-41^

We developed VISION-ACT with input from glaucoma patients and clinicians to ensure it is applicable to the glaucoma context and scalable to address psychological distress at the population level. We now aim to evaluate the feasibility and acceptability of VISION-ACT in a single-arm pilot, and explore preliminary changes in psychological distress and related patient-reported outcomes, including vision- and health-related QoL, psychological flexibility, and self-efficacy for symptom management. Our findings will inform the potential for a larger-scale, fully powered efficacy trial of VISION-ACT.

## METHOD

### Participants

This study was approved by the Duke University Institutional Review Board (IRB#: Pro00108155), registered on ClinicalTrials.gov (NCT06053307), and adhered to the tenets of the Declaration of Helsinki and all federal and state laws. Participants were recruited from the Duke Eye Center. Study enrollment occurred between April 2025 and December 2025. Participants inclusion criteria included: 1) ≥18 years old, 2) a diagnosis of mild, moderate, or severe glaucoma based on diagnosis codes (e.g., POAG), 3) a visual field within the past year at the Duke Eye Center, 4) prescribed pressure lowering eye drop medication, 5) at least mild (≥3) self-reported distress on the 4-item Patient Health Questionnaire (PHQ-4),^42^ 6) committed to engage in a 6-week mobile app behavioral intervention, and 7) able to understand, speak, and read English. Exclusion criteria included: 1) borderline glaucoma or glaucoma suspect, 2) glaucoma surgery in the past month (e.g., trabeculectomy, glaucoma drainger device/tube), 3) visual acuity of worse than 20/70 in the better seeing eye, 4) medical (e.g., macular degeneration, cognitive impairment) and/or psychiatric (e.g., psychosis) condition indicated by provder or electronic health record (EHR) review contraindicating safe participation.

### Procedure

Recruitment procedures complied with HIPAA guidelines. Potentially eligible patients were identified by EHR review and emailed a letter introducing the study. The introductory letter included an opt-out option, as well as a link to complete eligibility screening if interested. Study staff called patients that did not self-screen to describe the study purpose, procedures, risk and benefits, and provide potential participants with an opportunity to ask questions. Interested patients completed a brief screening interview via telephone to assess eligibility. If eligible and interested, patients electronically signed an IRB-approved consent form using Research Electronic Data Capture (REDCap). After consent, participants completed a self-report assessment via REDCap (i.e., baseline [A1]) and a telephone tutorial session with study staff involving app registration and orientation to the app format and its features. Follow-up assessments occurred at post-intervention (A2, primary endpoint) and 1-month post-intervention (A3). Participants were compensated $40 for each completed assessment. At the post-intervention (A2) timepoint, participants were invited to complete an optional exit interview, for which they were compensated an additional $30.

### Intervention

VISION-ACT is an asynchronous mobile app intervention developed by glaucoma physicians and clinical health psychologists with expertise in Acceptance and Commitment Therapy (ACT) for managing chronic disease symptoms. Aligning with the essential components of ACT,^26^ the VISION-ACT protocol was comprised of 6 modules addressing: 1) Being Present, 2) Cognitive Defusion, 3) Acceptance, 4) Self as Context, 5) Values, and 6) Committed Action (Figure 1). Modules were designed to be completed in the participant’s own environment, and at their convenience, weekly over the course of approximately six weeks. Each module included brief didactic (*Discover*) and skills (*Do*) videos incorporating text, visual, and audio content tailored to the glaucoma experience. For example, participants were prompted to use cognitive diffusion skills (i.e., Leaves on a Stream) to identify and let go of distressing thoughts related to their glaucoma, and use acceptance techniques (e.g., Slow Belly Breath) to acknowledge and be present with fears about disease progression. Strategies to adapt values-driven activities based on current level of visual functioning were also discussed. In-app activities were included to enhance engagement and reflection on skill practice (e.g., journaling). Participants were also provided with, and encouraged to complete paper worksheets to complement skill presentation. Text reminders and push notifications were used to further strengthen engagement. Figure 1 outlines VISION-ACT content and app features.

**Figure 1.**
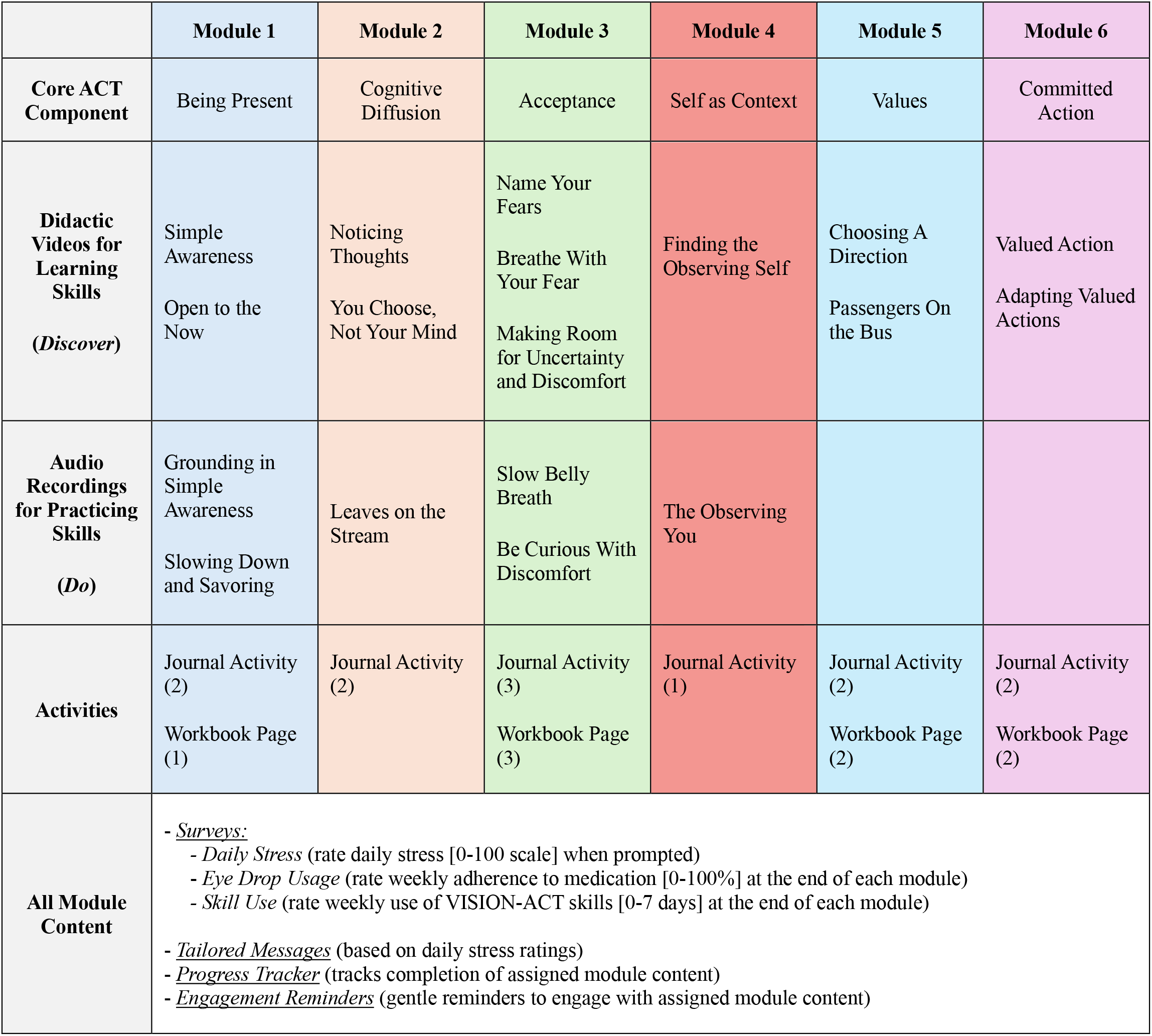
VISION-ACT Mobile App Content and Features

Participants’ stress level was assessed daily with a single-item survey in the app (“On a scale of 0 to 100, where 0 means no stress and 100 means extreme stress, how stressed are you feeling right now?”). Based on the participant’s response, tailored messages were sent via text with general encouragement for lower stress ratings (e.g., “Looks like a good day so far. Keep it up!”) and more directive recommendations for skill practice and app engagement for higher stress ratings (e.g., Can you use your favorite VISION-ACT skill now to help you reset?”).

VISION-ACT was hosted by Pattern Health (Greenlight Health Data Solutions Inc.) and downloaded for free by participants through iOS or Android on a smartphone device.

### Measures

#### Demographics and Clinical Characteristics

Demographic variables self-reported at baseline included age (in years), race (White, Black/African American, other), sex (female, male), ethnicity (Hispanic/Latino, non-Hispanic/Latino), marital status (single, married), highest level of education attained (high school or lower, greater than high school), income level (less than $100k/year, more than $100k/year), and employment status (retired, employed, disability, other). Clinical data was collected by manual EHR review. Eye-specific clinical data used in this study included intraocular pressure (mm Hg), visual acuity (LogMAR), and standard automated perimetry mean deviation (SAP MD, dB). Values for both the better eye (i.e., smaller values of IOP and LogMAR, higher values of SAP MD) and the worse eye (i.e., larger values of IOP and LogMAR, and lower values of SAP MD), as well as the average value, were collected.

#### Feasibility and Acceptability

A priori feasibility and acceptability benchmarks were based on behavioral intervention development guidelines and our prior feasibility work.^43-45^ Feasibility benchmarks were 1) target accrual (n=25) in 12 months and 2) <20% attrition at post-intervention (A2; primary endpoint). Acceptability benchmarks were 1) ≥75% of participants reporting use of skills/ideas from the intervention at A2 (“Overall, in the last week, how often have you used the skills or ideas presented in the VISION-ACT program?” Response Options: Not at All, One Time, A Few Days, Several Days, Almost Every Day, and Every Day) and 2) ≥80% of participants reporting satisfaction at post-intervention on the Client Satisfaction Questionnaire^46^ (i.e., 8-item CSQ mean ≥3.00/4.00).

#### Patient-Reported Outcomes

Psychological distress was assessed by the 1) Hospital Anxiety and Depression Scale (HADS)^47^ and 2) Subjective Units of Distress Scale (SUDS).^48^ The HADS has 7 items that assess anxiety symptoms (e.g., worry, tension) and 7 items that assess depressive symptoms (e.g., mood, interest in activities). Items are summed to yield anxiety (HADS-A) and/or depression (HADS-D) subscales, or an overall total score ranging from 0-42 with higher scores indicating higher distress. The SUDS is a single-item visual analog scale ranging from 0 (no distress) to 100 (extreme distress), and participants were asked to rate their distress “right now,” at the time of the assessment

Vision-related and health-related QoL were assessed by the National Eye Institute Visual Functioning Questionnaire^49^ (NEI-VFQ, 9-item, Range: 0-100, higher values indicate higher vision-related QoL) and the first item of the CDC Health-Related Quality of Life Questionnaire^50^ (1 item, rate general health from “excellent” to “poor”). Measures of psychological flexibility (Acceptance and Action Questionnaire [AAQ-2],^51^ 7-item, Range: 0-49, higher values indicate lower psychological flexibility), disease acceptance (Acceptance of Illness Scale [AIS],^52,53^ 8-item, Range: 8-40, higher scores indicate higher illness acceptance), self-efficacy for chronic disease management (Self-Efficacy for Managing Chronic Disease Scale [SE-6],^54^ 6-item, Range: 6-60, higher values indicate higher self-efficacy), mindfulness (Cognitive and Affective Mindfulness Scale-Revised [CAMS-R],^55^ 8-item, Range: 10-40, higher values indicate higher mindfulness), and social support (Medical Outcomes Study Social Support Survey [MOS-8],^56^ 8-item, Range: 8-40, higher values indicate higher perceived social support) were also assessed. Patient-reported outcome measures were selected based on brevity and strong psychometric properties in populations with chronic disease.

### Patient Perspectives

#### Preferences, Barriers, and Facilitors

At post-intervention, participants were asked 28 items modeled from our past work^33^ assessing preferences regarding referral to VISION-ACT (e.g., text, email) and content within the app (e.g., relaxation recordings, skills videos), as well as potential barriers (e.g., no smart phone, time commitment) and facilitators (e.g., user instruction, clinician involvement) for indivduals using VISION-ACT in the future. Items were answered on an 11-point Likert scale ranging from 0 (“Low Preference/Barrier/Facilitator”) to 10 (“High Preference/Barrier/Facilitator”).

#### Exit Interview

At post-intervention, participants were invited to compete an optional exit interview to share their feedback about the VISION-ACT protocol. Interviews were conducted by the study coordinator via telephone and lasted approximately 30 minutes. Field notes were used to record participant feedback, which was summarized and organized by interview question: 1) “What were your overall impressions of the VISION-ACT program?”, 2) “What did you like about VISION-ACT?”, 3) “What did you not like about VISION-ACT?”, and 4) “What could we do better?”.

### Statistical Analysis

Descriptive statistics were presented for baseline characteristics as mean and standard deviation (SD) for continuous variables and as counts and percentages for categorical variables. For each patient-reported outcome, we fit a linear mixed-effects model with a fixed effect for assessment (A1 as the reference category) and a random intercept for participant to account for within-subject correlation across assessments. Estimated marginal means are reported for each outcome at each assessment (A1, A2, A3) along with corresponding 95% confidence intervals (CI). Pairwise mean differences between assessments and corresponding 95% CI were also estimated from these models. Survey responses from related to preferences, facilitators, and barriers were summarized as median and interquartile range (IQR). All analyses were performed using R Statistical Software (v4.4.1; R Core Team 2024).

## RESULTS

### Study Flow

Study staff called 174 patients. Of those called, 125 were reached to introduce the study (n=49 could not be reached by phone and/or email). Fifty-three patients (53/125, 42%) declined screening; primary reasons included not being interested in research participation (n=34) and being too busy (n=7). Seventy-two patients (72/125, 58%) were screened for eligibility via telephone, and 30 met eligibility criteria (30/72, 42%). Screen failures were primarily due to not reporting enough distress (n=34) or having another visual disorder (n=3). Of the 30 patients that met eligibility criteria, 28 consented (28/30, 93%) and were enrolled in the study. Twenty-six participants started the VISION-ACT intervention; two participants withdrew prior to starting the intervention due to competing demands/time commitment. See Figure 2 for the CONSORT diagram.

**Figure 2.**
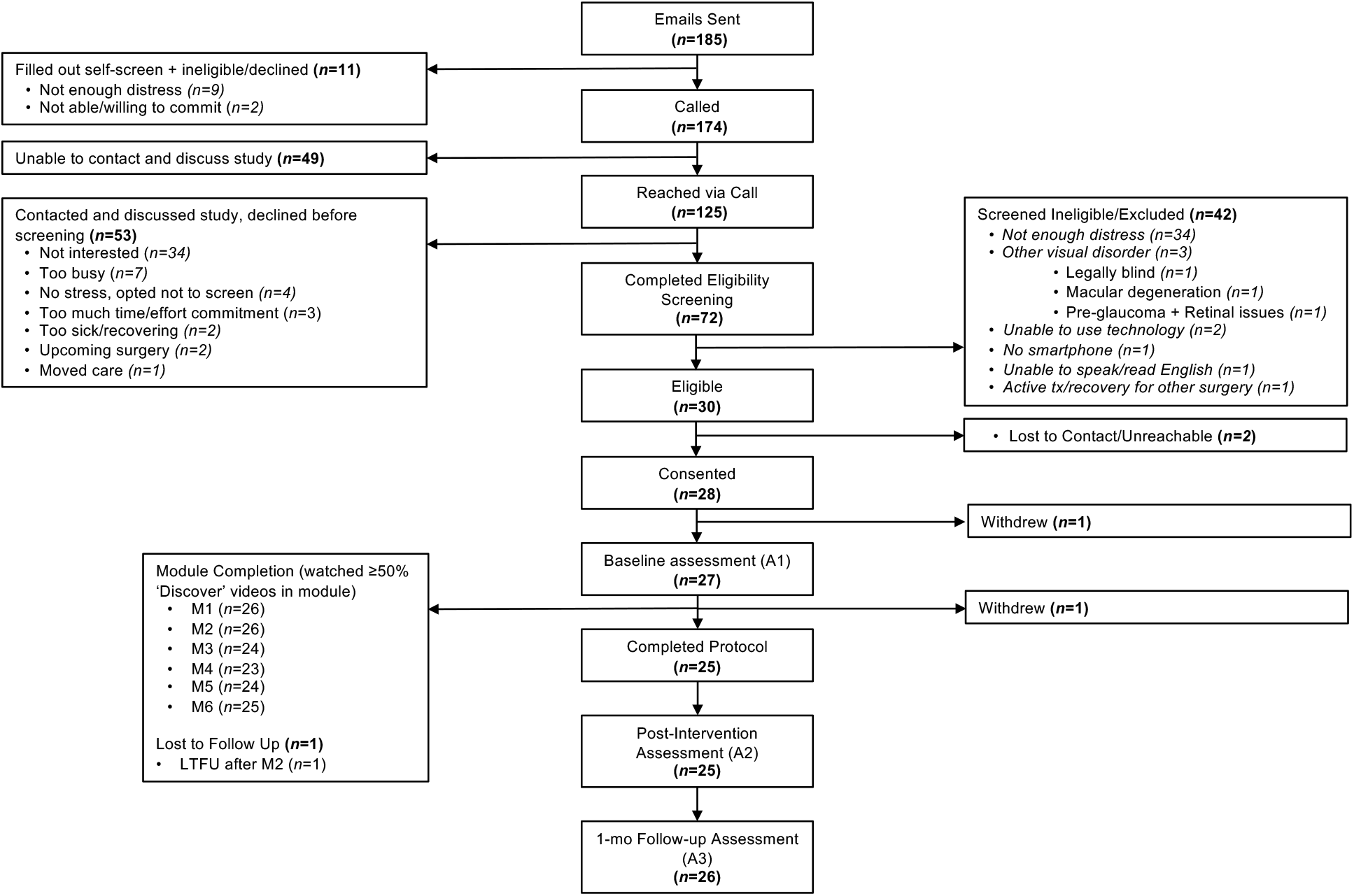
Study CONSORT diagram.

### Participant Characteristrics

Average age was 66.75 (SD=8.14) and participants were mostly White (58%) and female (65%). The majority of participants were married (77%) and retired (62%), and reported Some College or more (96%). Average IOP (mm Hg), visual acuity (LogMAR), and standard automated perimetry mean deviation (dB) for the better eye were 12.65 (SD=3.20), 0.19 (SD=0.24), and -3.92 (SD=6.03), respectively. Table 1 presents demographic and clinical variables for the sample of participants that started the VISION-ACT intervention (n=26).

### Feasibility

The original recruitment goal (n=25) was met in 6 months, well ahead of the 12-month accrual benchmark. Three additional participants beyond our goal were recruited due to interest in the study, bringing total number of consented participants to 28. Of the 26 participants who ultimately started the intervention, only 1 (3.85%) did not complete the post-intervention assessment (A2; primary endpoint); thus, our A2 attrition benchmark (<20%) was met.

### Acceptability

At A2, all participants (100%) reported use of skills and ideas from VISION-ACT in the past week, exceeding the ≥75% benchmark. Specifically, 12% reported use of intervention skills and ideas “Every Day,” 8% “Almost Every Day,” 36% “Several Days,” 40% “A Few Days,” and 4% “One Time.” There were no participants that reported using skills/ideas “Not at All.” Adherence to VISION-ACT was strong, with 23 participants completing all six modules (23/26, 88.5%). Only 3 participants missed some, but not all, module content. The proportion of participants that reported an average satisfaction score of ≥3.00/4.00 was 76%, just below the ≥80% benchmark. However, the average satisfaction score at A2 was above 3.00 at 3.27 (SD=0.57).

### Patient-Reported Outcomes

Participants improved from A1 to A2 on psychological distress (Table 2). Total HADS psychological distress score decreased from A1 (estimated mean [M]=13.88, 95% CI [11.57, 16.20]) to A2 (M=11.21, 95% CI [8.88, 13.54]), as did HADS-Anxiety (A1 M=8.38, 95% CI [6.81, 9.96]; A2 M=7.27, 95% CI [5.69, 8.85]) and HADS-Depression subscales (A1 M=5.50, 95% CI [4.46, 6.54]; A2 M=3.94, 95% CI [2.90, 4.99]). Subjective Units of Distress scores also decreased from A1 (M=35.54, 95% CI [26.98, 44.10]) to A2 (M=26.46, 95% CI [17.81, 35.12]). Vision-related quality of life increased from A1 (M=73.12, 95% CI [68.36, 77.88]) to A2 (M=79.88, 95% CI [75.09, 84.66]). The proportion of participants reporting “Very Good” and “Excellent” general health was relatively consistent at A1 (34.6%) and A2 (36%), although increased to 46.1% at A3.

**Table 2.** Estimated marginal means and mean differences for patient-reported outcomes across the baseline (A1), post-intervention (A2), and 1-month post-intervention (A3) assessments. Values are reported as model-based estimated means and pairwise mean differences with corresponding 95% confidence intervals from linear mixed models.

| Measure | A1 | A2 | A3 | A1-A2 | A1-A3 | A2-A3 |
| --- | --- | --- | --- | --- | --- | --- |
| HADS | 13.88 (11.57, 16.20) | 11.21 (8.88, 13.54) | 9.38 (7.07, 11.70) | 2.67 (1.05, 4.29) | 4.50 (2.90, 6.10) | 1.83 (0.21, 3.45) |
| HADS-A | 8.38 (6.81, 9.96) | 7.27 (5.69, 8.85) | 5.81 (4.23, 7.38) | 1.11 (0.01, 2.22) | 2.58 (1.49, 3.66) | 1.46 (0.36, 2.57) |
| HADS-D | 5.50 (4.46, 6.54) | 3.94 (2.90, 4.99) | 3.58 (2.54, 4.61) | 1.56 (0.70, 2.41) | 1.92 (1.08, 2.76) | 0.37 (-0.49, 1.22) |
| SUDS | 35.54 (26.98, 44.10) | 26.46 (17.81, 35.12) | 26.23 (17.67, 34.79) | 9.08 (0.93, 17.22) | 9.31 (1.27, 17.35) | 0.23 (-7.92, 8.38) |
| NEI-VFQ-9 | 73.12 (68.36, 77.88) | 79.88 (75.09, 84.66) | 79.40 (74.64, 84.17) | -6.76 (-9.45, -4.06) | -6.28 (-8.94, -3.63) | 0.47 (-2.22, 3.17) |
| AAQ-2 | 17.42 (14.19, 20.65) | 14.61 (11.36, 17.86) | 14.31 (11.08, 17.54) | 2.81 (0.69, 4.94) | 3.12 (1.02, 5.21) | 0.30 (-1.82, 2.42) |
| AIS | 29.50 (26.37, 32.63) | 30.65 (27.48, 33.81) | 33.08 (29.95, 36.20) | -1.15 (-4.29, 2.00) | -3.58 (-6.68, -0.47) | -2.43 (-5.57, 0.71) |
| SE-6 | 6.87 (6.08, 7.66) | 7.79 (6.99, 8.58) | 8.25 (7.46, 9.04) | -0.92 (-1.53, -0.30) | -1.38 (-1.99, -0.77) | -0.46 (-1.08, 0.16) |
| CAMS-R | 27.69 (25.08, 30.30) | 30.21 (27.59, 32.84) | 30.85 (28.24, 33.45) | -2.52 (-4.41, -0.63) | -3.15 (-5.02, -1.29) | -0.63 (-2.52, 1.26) |
| MOS-8 | 31.35 (27.84, 34.85) | 32.05 (28.54, 35.57) | 33.15 (29.65, 36.66) | -0.71 (-2.34, 0.92) | -1.81 (-3.41, -0.20) | -1.10 (-2.73, 0.53) |

HADS, Hospital Anxiety and Depression Scale; HADS-A, Anxiety Subscale; HADS-D, Depression Subscale, SUDS, Subjective Units of Distress, NEI-VFQ-9, National Eye Institute Visual Function Questionnaire; AAQ-2, Acceptance and Action Questionnaire; AIS, Acceptance of Illness Scale; SE-6, Self-Efficacy for Managing Chronic Disease Scale; CAMS-R, Cognitive and Affective Mindfulness Scale-Revised; MOS-8, Medical Outcomes Study Social Support Survey.

Outcome change patterns were favorable and in expected directions for all other patient-reported outcomes (psychological flexibility, disease acceptance, self-efficacy for chronic disease management, mindfulness, and social support), and improvements continued out to A3. Estimated marginal means, pairwise changes, and corresponding 95% CI and are shown in Table 2 and visualizations of change patterns are shown in Figure 3.

**Figure 3.**
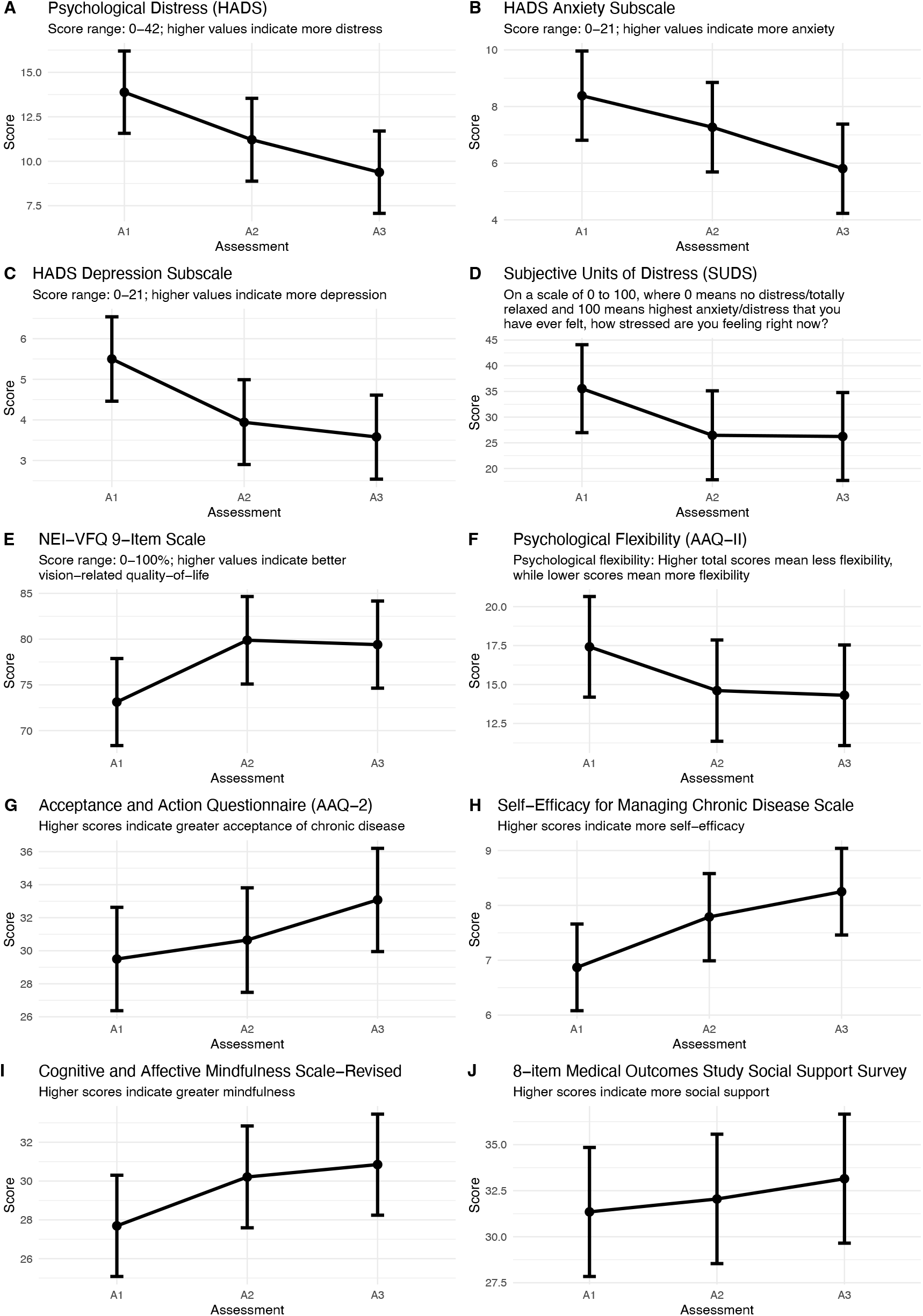
Estimated marginal means with corresponding 95% confidence intervals for patient-reported outcomes across the baseline (A1), end of study (A2), and one-month post study (A3) assessments. NEI-VFQ, National Eye Institute Visual Function Questionnaire.

### Patient Perspectives

#### Preferences, Barriers, and Facilitors

Figure 4 shows the median and interquartile range (IQR) for items assessing preferences for referral to VISION-ACT and intervention content, as well as participants’ impressions of barriers and facilitators to VISION-ACT use. Participants most preferred being referred to VISION-ACT through a direct text message to their phone. Content preference items indicated that participants wanted more audio relaxation recordings and increased involvement of a glaucoma clinician. Additionally, participants endorsed wanting more intervention modules, as well as more videos and patient stories reviewing use of, and experience with coping skills. The highest rated barrier to someone using VISION-ACT in the future was not having a smart phone. The highest rated facilitator was referral from a glaucoma clinician.

**Figure 4.**
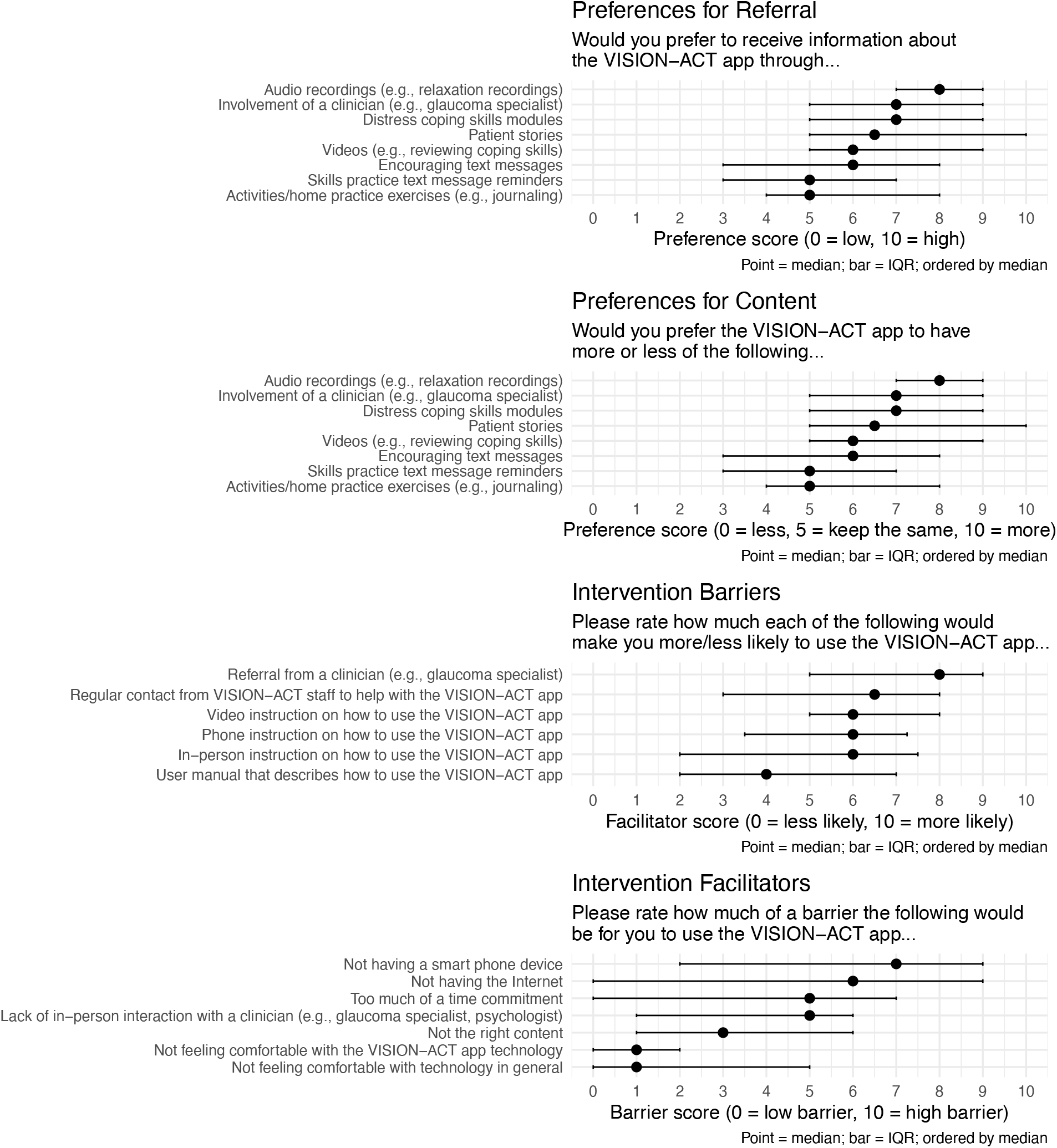
Preferences for Intervention Referral and Content, and Intervention Barriers and Facilitators. Results are summarized as median and interquartile range (IQR).

#### Exit Interview

Twenty-two participants agreed to complete the exit interview. Overall impressions of VISION-ACT were positive, with participants sharing that the program helped them learn relaxation strategies, gain insight, and receive emotional support. VISION-ACT was described as “calming,” “enlightening,” and “meaningful.” A subset of skills were particularly well-liked and included Leaves on a Stream, Slow Belly Breath, and Valued Actions. These skills were noted to be “accessible,” “soothing,” and broadly applicable to both glaucoma-related and general life stressors. Participants also shared that they appreciated the self-paced structure of the app and need for minimal staff contact.

Areas for improvement centered mostly on app features and usability. Reminders from the app to use VISION-ACT skills were reported to be helpful but also a source of frustration due large amount, high frequency, and random timing. Worksheets that accompanied app modules had mixed reviews, with some finding them useful while others felt they were a burden (i.e., those preferring mental reflection). Suggestions for improvement included 1) introducing VISION-ACT earlier in the glaucoma experience (e.g., at diagnosis), 2) fewer app messages/reminders, with more encouraging wording, and 3) more clear schedule of required videos.

## DISCUSSION

In this study, we pilot tested the feasibility and acceptability of a standalone, mobile app psychosocial intervention (VISON-ACT) for patients diagnosed with glaucoma and reporting psychological distress. VISION-ACT exceeded all feasibility benchmarks. Acceptability was strong and exploratory estimates suggest that participants improved on psychological distress and all other patient-reported outcomes from baseline to post- and 1-month post-intervention.

At present, there are no evidence-based, psychosocial interventions addressing psychological distress during the glaucoma experience. This is a critical gap in the field, given psychological distress is a central concern for glaucoma patients and related to key clinical outcomes (e.g., treatment adherence, disease progression).^6,8,11^ Formative, early-stage intervention development work is urgently needed to carefully develop and test psychosocial protocols that are applicable and beneficial to glaucoma patients and hold potential for widespread implementation.

VISION-ACT addresses these objectives, as the first ACT-based intervention tailored specifically to glaucoma patients with psychological distress, and delivered asynchronously via a mobile app. VISION-ACT demonstrated impressive feasibility and acceptability. We met our recruitment goal (N=25) 6 months ahead of schedule, and surpassed our goal by enrolling three additional participants who expressed interest in the study. Nearly 60% of contacted patients were agreeable to eligibility screening, and of those, 42% met inclusion criteria. Strikingly, 93% of those screened, were consented and enrolled. Together, these data suggest that recruitment procedures are feasible and the protocol is appealing to patients with glaucoma. Retention was excellent, as only 1 participant did not complete the post-intervention assessment (primary endpoint). This low attrition rate is especially remarkable for an asynchronous intervention with minimal staff contact.

Acceptability data are also promising. At post-intervention, all participants reported use of skills and ideas from VISION-ACT in the past week. This was well above our prespecified benchmark (≥75%) and suggests that participants found the intervention content to be highly relevant. Relatedly, adherence was excellent with 88.5% of participants completing all six VISION-ACT modules. This adherence rate is notable for an entirely self-paced intervention without any therapist involvement. Indeed, similar standalone internet or mobile app-delivered interventions in other chronic disease populations typically report adherence rates at or below 70%.^37,57,58^ While the proportion of participants (76%) reporting an average satisfaction score of ≥3.00/4.00 was slightly below the ≥80% benchmark, mean satisfaction was 3.27, suggesting a largely favorable review.

Data on patient perspectives and qualitative feedback obtained during exit interviews provide useful context to acceptability findings. Valued aspects of the intervention were its self-paced format, widely applicable mindfulness and acceptance skills (e.g., Leaves on a Stream, Slow Belly Breath), and strategies for identifying behaviors that are aligned with values. Key areas of improvements that may enhance acceptability for next iterations of VISION-ACT include reducing the number and frequency of app messages and reminders, and clarifying the schedule of required videos. Additionally, participants shared a desire for more intervention modules and audio relaxation recordings. It is possible that incorporating booster modules after the primary intervention period may serve as a welcome reminder of skills and ultimately help maintain treatment gains. Finally, a recurring theme from participant feedback was the importance of involving the glaucoma clinician. Participants consistently shared that endorsement of VISION-ACT from their clinicians was critical and would enhance motivation to engage with the program.

This was a single-arm pilot study with a primary aim of assessing initial feasibility and acceptability of the VISION-ACT protocol. Still, pre-post change patterns on patient-reported outcomes were explored, and showed that participants improved on psychological distress. Estimated mean total HADS score, and anxiety and depression subscale scores, all decreased from baseline to post-intervention; HADS scores further decreased out to the 1-month post-intervention timepoint. A similar pattern was observed for the Subjective Units of Distress scale, which reduced by nearly 10 points from baseline to post-intervention. All other patient-reported outcomes (vision- and health-related quality of life, psychological flexibility, disease acceptance, self-efficacy for disease management, mindfulness, social support) also improved across baseline, post-intervention, and 1-month post-intervention assessments. Therapist-led, ACT interventions have demonstrated small-to-medium effects on psychological distress and quality of life in other chronic disease populations, and mobile app delivered ACT protocols, specifically, are beginning to show promise.^24,25,35^ This is the first time a mobile app-delivered ACT intervention has been examined in a glaucoma sample. Results presented here are preliminary and should be interpreted with caution given the single-arm design and small sample size. Nevertheless, it is encouraging that psychological distress and related patient-reported outcomes improved in the expected direction, indicating that further testing is warranted for this patient population.

This study was not a randomized trial, and the sample was small. Thus, we were not powered for hypothesis or efficacy testing. This is a limitation of pilot work. The next step efficacy trial will include a larger sample that provides adequate power for testing VISION-ACT against an time and attention-matched control condition. Participants were enrolled from a large academic medical center and mostly White, female, and highly educated. As such, findings may not be representative of the diversity seen in the glaucoma population, generally. Participants were at different stages of their disease, with some being recently diagnosed and other being years out from diagnosis. Severity of psychological distress is likely to vary based on phase of disease management (i.e., diagnosis, surveillance), and this may impact engagement with VISION-ACT, as well as its perceived usefulness. Finally, clinical outcomes (e.g., intraocular pressure variability) were not assessed longitudinally, despite observed correlations to psychological distress. Future work should endeavor to examine how completion of VISION-ACT might improve psychosocial and clinical variables, and how these improvements may relate to each other.

### Clinical Implications and Future Directions

This study was the first in its field to rigorously develop and pilot test an ACT-based, psychosocial intervention tailored to the challenging glaucoma experience, using a scalable mobile app delivery format. Qualitative feedback will inform further refinement to the protocol. Specifically, app messaging will be reduced and the schedule of module videos will be clarified. Additionally, involvement of glaucoma clinicians will be thoughtfully increased through educational videos about glaucoma and its treatment and direct referral to VISION-ACT from glaucoma physicians at the point of care. Robust feasibility and acceptability data seen here provide support a fully-powered, randomized trial to evaluate the efficacy of VISION-ACT for reducing psychological distress and improving related patient-reported and clinical outcomes. Ultimately, if results from this line of research are positive, our efforts will quickly shift to implementation to help more glaucoma patients receive empirically-supported, psychosocial support for the significant psychological burden of this chronic disease.

## Data Availability

All data produced in the present study are available upon reasonable request to the authors and will be made available online in a repository.

## ACKNOWLEDGMENTS

Research reported in this publication was supported by the National Eye Institute of the National Institutes of Health (Bethesda, Maryland) under Awards Number R00EY033027 (SIB), R01EY029885 (FAM), and R01EY036593 (FAM). The sponsor or funding organization had no role in the design or conduct of this research. The content is solely the responsibility of the authors and does not necessarily represent the official views of the National Institutes of Health.

## SUPPLEMENTARY TABLES

**Table S1.**
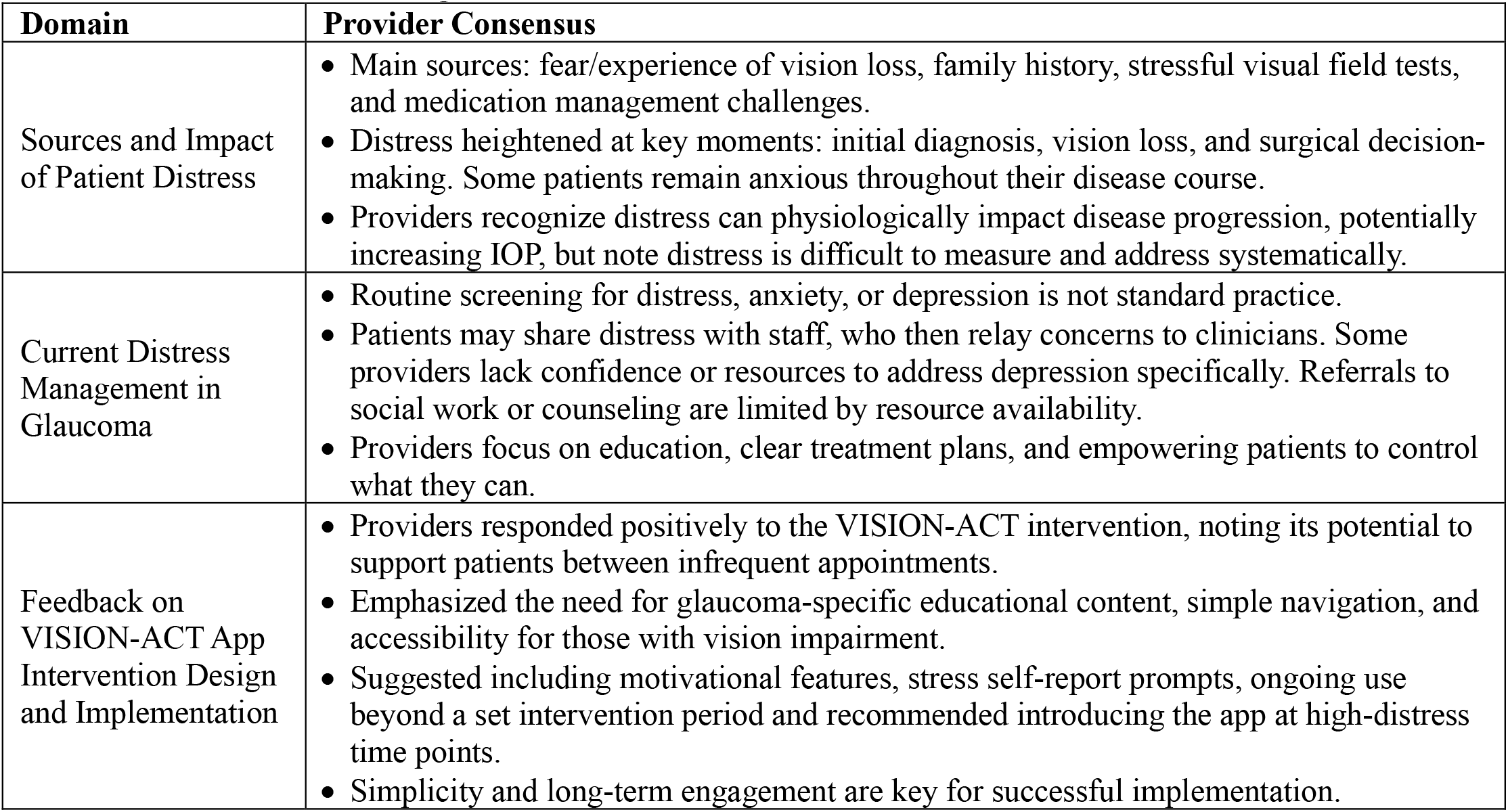
Qualitative findings from semi-structured clinician interviews.

**Table S2.** Qualitative findings from semi-structured patient interviews.

| Phase | Patient Consensus |
| --- | --- |
| Phase 1-3<br>(n=16) | • **Stress and Uncertainty:** Most experienced significant stress at diagnosis, especially those with symptoms, younger age, or with family history. Stress persisted with disease progression, vision loss, and uncertainty about the future.<br>• **Understanding and Management:** Understood glaucoma as a progressive, manageable disease requiring regular monitoring and medication. Adherence was high with side effects and complex regimens posing challenges for some.<br>• **Impact on Daily Life:** Vision loss affected work, independence, driving, and leisure. Some made major life adjustments; others reported minimal impact.<br>• **Coping Strategies:** Used faith, mindfulness, acceptance, positive attitude, support from family/providers, routines, and some unhealthy coping habits.<br>• **Communication with Providers:** Trust and clear communication with providers reduced stress. Most felt comfortable asking questions; value patient portals.<br>• **Medication Adherence:** Most consistent with drops, using reminders and routines. Side effects and multiple medications were barriers for a few.<br>• **Feedback on Intervention/App:** Patients valued education, reminders, stress management tools, and support for self-management. They wanted flexibility, accessible design, and opportunities for human contact or peer support. |
| Phase 4<br>User<br>Testing<br>(n=4) | • **Usability and Accessibility:** App generally easy to use, with clear instructions and logical layout. High contrast and large fonts were recommended for those with more vision loss. Minimal technical support needed, but in-person onboarding helpful for some.<br>• **Engagement and Content:** Initial engagement was high. Over time repetitive content reduced motivation. Enjoyed mindfulness and stress management features.<br>• **Navigation and Flexibility:** Users wanted more flexible module timing, the ability to revisit content, and personalized reminders (for all medications). Some found the task list overwhelming or module timing inflexible.<br>• **Perceived Value and Suggestions:** Patients appreciated the app's grounding effect during stress and want clearer links between app activities and glaucoma outcomes. Suggestions included more relatable stories, group support options, and enhanced personalization. Provider endorsement and repeated introduction at key disease milestones were seen as important for uptake. |

